# *GCH1* mutations in hereditary spastic paraplegia

**DOI:** 10.1101/2021.01.14.21249305

**Authors:** Parizad Varghaei, Grace Yoon, Mehrdad A Estiar, Simon Veyron, Etienne Leveille, Nicolas Dupre, Jean-François Trempe, Guy A Rouleau, Ziv Gan-Or

**Affiliations:** Division of Experimental Medicine, Department of Medicine, McGill University, Montreal, Quebec, Canada; Montreal Neurological Institute and Hospital, McGill University, Montreal, Quebec, Canada; Divisions of Neurology and Clinical and Metabolic Genetics, Department of Pediatrics, University of Toronto, The Hospital for Sick Children, Toronto, Canada; Department of Human Genetics, McGill University, Montreal, Quebec, Canada; Department of Pharmacology & Therapeutics and Centre de Recherche en Biologie Structurale - FRQS, McGill University, Montréal, Canada; Faculty of Medicine, McGill University, Montreal, QC, Canada; Axe Neurosciences, CHU de Québec-Université Laval, Québec, QC, Canada; Department of Medicine, Faculty of Medicine, Université Laval, Quebec City, QC, Canada; Department of Neurology and Neurosurgery, McGill University, Montreal, Quebec, Canada

**Author notes:** Correspondence to: Guy A. Rouleau, Director, Montréal Neurological Institute and hospital, Address: 3801, University Street, Office 636, Montréal, Québec H3A 2B4, Ziv Gan-Or, Department of Neurology and Neurosurgery, McGill University, 1033 Pine Avenue, West, Ludmer Pavilion, room 312, Montreal, QC, H3A 1A1.

**Keywords:** HSP, GCH1, Dystonia, Spastic paraplegia

## Abstract

*GCH1* mutations have been associated with dopa-responsive dystonia (DRD), Parkinson’s disease (PD) and tetrahydrobiopterin (BH_4_)-deficient hyperphenylalaninemia B. Recently, *GCH1* mutations have been reported in five patients with hereditary spastic paraplegia (HSP). Here, we analyzed a total of 400 HSP patients (291 families) from different centers across Canada by whole exome sequencing (WES). Three patients with heterozygous *GCH1* variants were identified: monozygotic twins with a p.(Ser77_Leu82del) variant, and a patient with a p.(Val205Glu) variant. The former variant is predicted to be likely pathogenic and the latter is pathogenic. The three patients presented with childhood-onset lower limb spasticity, hyperreflexia and abnormal plantar responses. One of the patients had diurnal fluctuations, and none had parkinsonism or dystonia. Phenotypic differences between the monozygotic twins were observed, who responded well to levodopa treatment. Pathway enrichment analysis suggested that GCH1 shares processes and pathways with other HSP-associated genes, and structural analysis of the variants indicated a disruptive effect. In conclusion, *GCH1* mutations may cause HSP; therefore, we suggest a levodopa trial in HSP patients and including *GCH1* in the screening panels of HSP genes. Clinical differences between monozygotic twins suggest that environmental factors, epigenetics, and stochasticity could play a role in the clinical presentation.

## Introduction

The guanosine triphosphate cyclohydrolase I gene (*GCH1*, [OMIM] *600225) encodes GTP cyclohydrolase 1, a biosynthetic enzyme which catalyzes the synthesis of tetrahydrobiopterin (BH_4_). BH_4_ is a co-factor of tyrosine hydroxylase, an enzyme involved in the synthesis of the neurotransmitters dopamine and monoamines.^1,2^ Due to its importance in the synthesis of these neurotransmitters, several neurological disorders may result due to GTP cyclohydrolase 1 deficiency.

Rare bi-allelic *GCH1* mutations may cause tetrahydrobiopterin (BH_4_)-deficient hyperphenylalaninemia B (HPABH4B, OMIM #233910),^3^ a rare disease which may include severe developmental delay, muscular hypotonia of the trunk, hypertonia of the extremities, seizures and other neurological symptoms.^4-6^ Rare heterozygous mutations in *GCH1* are known to cause autosomal dominant dopa-responsive dystonia (DRD), which typically presents with dystonia of the lower limbs in childhood, diurnal fluctuations and a substantial response to levodopa treatment.^7-10^ In recent years it has been shown that rare heterozygous *GCH1* mutations may also cause Parkinson’s disease (PD),^11-14^ and that common variants in the *GCH1* locus are associated with risk of PD.^14,15^

Two previous reports have implicated *GCH1* mutations in hereditary spastic paraplegia (HSP) patients,^16,17^ yet their role in HSP is still not conclusive. HSP is a general term for a group of inherited neurodegenerative disorders characterized by lower limb spasticity and weakness, with or without additional neurological symptoms.^18^ Herein, we report three additional HSP patients from two different families with heterozygous *GCH1* mutations, their phenotype and long-term follow-up.

## Materials and Methods

### Population

HSP patients (n=696) from 431 families were enrolled to CanHSP, a Canadian consortium for the study of HSP.^19^ The patients were recruited from eight major medical centers across Canada (Montreal, Toronto, Quebec, Calgary, Edmonton, Ottawa, Hamilton, and Vancouver). Data regarding diagnosis, recruitment, and the cohort have been previously described.^19^ All patients signed informed consent forms at enrollment and the study protocol was approved by the institutional review board.

### Genetic analysis

Genomic DNA has been extracted from peripheral blood, according to standard procedures.^20^ After diagnosis of a portion of the patients with panel sequencing, WES was performed on 400 patients from 291 families, using the Agilent SureSelect Human All Exon v4 kit for capture and targeted enrichment of the exome. Captured samples were sequenced in Illumina HiSeq 2000/2500/4000 system. Then, the sequence reads were aligned against the human genome (GRCh37 assembly) using Burrows-Wheeler Aligner (BWA).^21^ Variant calling and annotation were done using Genome Analysis ToolKit (GATK)^22^ and Annotate Variation (ANNOVAR).^23^

In order to rule out other genes known to be involved in HSP or in similar neurogenetic disorders in which spasticity is among the features, we analyzed the data using a list of 785 such genes (Supplementary Table 1). The nomenclature of *GCH1* variants is based on its full transcript NM_000161. We used MutationTaster, FATHMM, EIGEN, SIFT, CADD and REVEL for assessing the pathogenicity of the variants.^24^ Variants were classified according to the guidelines of American College of Medical Genetics and Genomics (ACMG) using VarSome.^25^

Domain prediction was performed using Interpro,^26^ and the reported mutations in PD, DRD and HSP were divided in two groups: within the main domain of the GCH1 protein (GTP_Cyclohydrolase_I_domain [IPR020602]) or outside of the main domain. Pearson Chi-square test was applied to determine whether an association between these two categorical variables (disease type and mutation location) exists. Gene Ontology (GO) enrichment analysis of *GCH1* with a list of known HSP associated genes (Supplementary table 2) was carried out using the g:Profiler,^27^ with Benjamini-Hochberg adjusted *p*-values for statistical significance set at < 0.05.

### In silico structural analysis

The atomic coordinates of human GCH1 bound to Zn^2+^ were downloaded from the Protein Data Bank (ID 1FB1). The effect induced by each mutation was evaluated using the “mutagenesis” toolbox in PyMol v. 2.4.0.

## Results

### Identification of *GCH1* mutations in HSP patients

Heterozygous *GCH1* mutations were identified in 3 HSP patients (representing 0.75% of HSP cases and 0.69% of HSP families with available WES data), including monozygotic twins from Family 1 and a single patient from Family 2. In family 1, following filtration, a novel heterozygous in-frame deletion in *GCH1*, c.229_246del: p.(Ser77_Leu82del), remained the only potential explanation for the phenotype of the twins. This variant was predicted to be likely pathogenic (Supplementary table 3) and was not reported in gnomAD.^28^ The affected twins were adopted, and family history or DNA from genetic relatives are not available. In family 2, another *GCH1* variant was identified, c.614T>A: p.(Val205Glu), which is not reported in gnomAD, and has been analyzed in two functional studies. These studies have suggested a significant decrease in the activity ^29^ and a strong effect on the function of the protein. ^30^ In VarSome, this variant has been classified as likely pathogenic, however, when taking into account the PS3 criterion (which concerns functional studies), the verdict changes to pathogenic.

Sanger sequencing of DNA samples from the proband’s parents indicated that the father carried the heterozygous mutation, while the mother had wild-type alleles. According to the proband, the father had no symptoms, but he was not seen by a neurologist. The variant is predicted to be deleterious by MutationTaster, FATHMM, EIGEN, SIFT, CADD (score of 26, within the top 0.5% of variants predicted to be most deleterious) and REVEL. Both variants occurred in the GTP_Cyclohydrolase_I_domain (IPR020602) which plays an important role in catalyzing the biosynthesis of formic acid and dihydroneopterin triphosphate from GTP. ^31^ All variants were validated by Sanger sequencing.

### Structural analysis of GCH1 pathogenic mutations

The structure of human GCH1 was solved by X-ray crystallography by Auerbach and colleagues.^32^ The structure reveals a toroid-shaped D5-symmetric homodecameric assembly, with the catalytic sites located at the interface of three adjacent subunits (Figure 1A). By homology with its *E. coli* ortholog, the catalytic site is defined as Cys141, His210 and Cys212 from one subunit, Ser166 from a second subunit and helix_92-104_ from a third subunit^33,34^ (Figure 1B). To investigate the impact of the degenerative mutations on the structure, we performed *in silico* mutagenesis and evaluated the effect on the surrounding residues.

**Figure 1.**
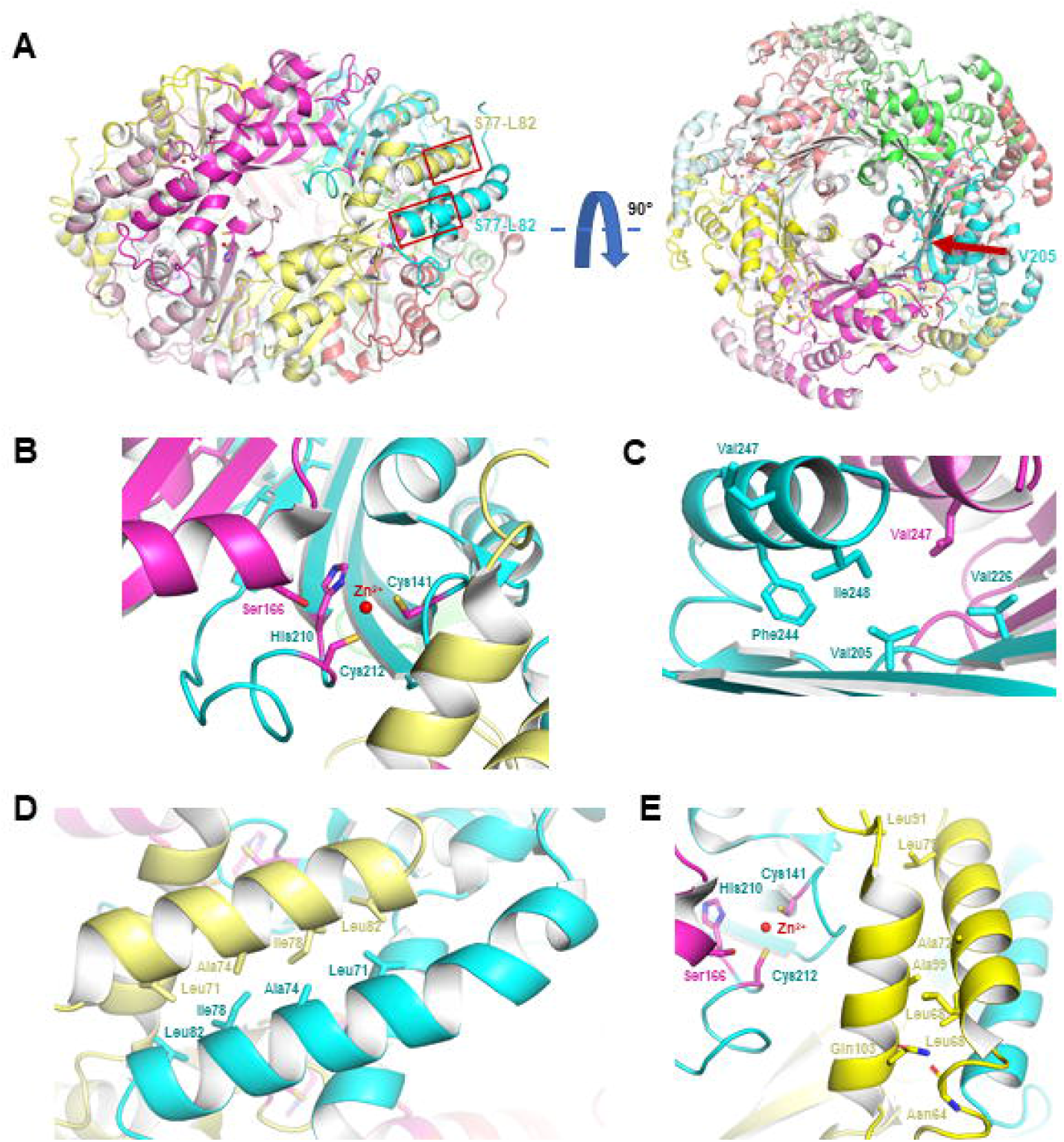
Structural analysis of GCH1 (protein database bank (PDB) code: 1FB1). A: Human GCH1 forms an homodecameric assembly. The Ser77-Leu82 region of subunit B (cyan) and D (pale yellow) are squared (left panel) and Val205 of subunit B is indicated with an arrow (right panel). B: The interface between subunit A (pink), B and D form an active site. The catalytic Cys141, Ser166, His210 and Cys212 are colored in magenta. C: Subunit B Val205 interacts with Phe244, Val226 and Ile248 of the same subunit, and Val247 of subunit A. D: Leu71, Ala74, Ile78 and Leu82 from subunit B and D form a hydrophobic-type interaction. E: Subunit D helix60-82 stabilize helix92-104 through the backbone OH of Asn64 and Gln103 sidechain and hydrophobic residues Leu68, Leu71, Ala72, Leu79, Leu91, Ala99.

Val205 is located on a β-sheet (strand_202-209_) and its sidechain points toward the core of the toroid, facing the C-terminal helices of two adjacent subunits. Its sidechain form hydrophobic interactions with Phe244, Val226 and Ile248 from the same subunit, and Val247 from the adjacent subunit, causing a leucine-zipper network of interactions, as valine, isoleucine and leucine have the same coupling energy^35^ (Figure 1C). The HSP-associated mutation of Val205 to the polar residue glutamate would therefore strongly disrupt these interactions. Therefore, the Val205Glu mutation will likely destabilize the entire β-sheet and possibly the entire domain, hence preventing its oligomerization and, *in fine*, its activity. By contrast, the DRD-associated mutation of Val205 into Gly likely results in a milder destabilization of this zipper, weakening the stabilization of the C-terminal helix on the inside of the toroid on one hand, and destabilizing the interaction between adjacent subunits. In addition, the Val205Glu would also induce more flexibility in the protein backbone as the absence of sidechain in glycine induce another degree of flexibility compared to others amino acids. Finally, several other mutations on the same β-sheet or on the facing helix have been reported to be causing DRD, supporting the hypothesis that disturbing this area is pathogenic.^36^

Residues Ser77 to Leu82 are located on a helix located on the outside of the toroid. This helix is involved in a homotypic interaction between two subunits on different halves of the toroid, through hydrophobic residues Leu71, Ala74, Ile78 and Leu82 from both helices (Figure 1D). As the two helices are anti-parallel, the Ser77_Leu82 deletion will abolish the interaction. As a result, this mutation will likely prevent the assembly of the 2 halves, either leading to homopentamers or to unstable monomers. In addition, helix_60-82_ interacts with helix_92-104_ near the catalytic site through the backbone OH of Asn64 and Gln103 sidechain and hydrophobic residues Leu68, Leu71, Ala72, Leu79, Leu91, Ala99 (Figure 1E). The Ser77_Leu82 deletion will shorten helix_60-82_, shifting its residues and likely disturbing its interaction with helix_92-104_ and disrupting its positioning as part of the catalytic site. Either way, this deletion will diminish the probability of formation of the active site. DRD-associated mutations Leu71Gln, Ala74Val and Leu79Pro have also been reported in this helix, supporting the hypothesis that disrupting the interaction between those helices is pathogenic, and that the degree of disruption of GCH1 leads to different pathologies.

In conclusion, the mutations Val205Glu and Ser77_Leu82del are probably affecting the proper homodecamerization of human GCH1, preventing the assembly of the three subunit-formed active sites. If Val205Glu seems to destabilize the proper folding of each monomer, Ser77_Leu82del probably prevents the polymerization of the homodecamer. Overall, those mutations appear to be “loss-of-function” but it would be interesting to investigate whether helix_92-104_ and the “inside of the toroid” have solely architectural roles, or function as well in catalysis.

### Pathway enrichment analysis

Pathway enrichment analysis suggests that GCH1 shares similar processes and pathways with other HSP-associated genes, including: hydrolase activity, nucleoside-triphosphatase activity, small molecule binding, catalytic activity, pyrophosphatase activity, organic acid biosynthetic process, carboxylic acid biosynthetic process, oxoacid metabolic process, regulation of anatomical structure size and small molecule biosynthetic process (Supplementary Table 4).

### Characteristics of HSP patients with *GCH1* mutations

#### Family 1

Cases A and B are monozygotic twins conceived naturally to a healthy mother, and born at 31.5 weeks of gestation. Both twins were adopted and live with their adoptive father and mother. Family history was reportedly negative for genetic or metabolic disorders, congenital malformations, neurological disorders, children with spasticity, seizure or mental challenges, history of other psychiatric disorders or autism.

##### Twin A clinical presentation

During the first years of life, twin A achieved developmental milestones at the appropriate time, and there was no early history of speech delay, vision or hearing problems. At an age range between 0 to 5 years of age, twin A started to show abnormality in their gait. Their symptoms progressed slowly over the years, and they had multiple episodes of falling every week, and later required aid for ambulating. They could sit normally, had normal bladder control and attended a regular class with grades above average. On physical exam between the ages 10-15 they had abnormal gait, bilateral spasticity of the lower limbs, abnormal posture of the feet, history of frequent falls, and use of walking aids most of the time. Their neurological exam showed spasticity of lower limbs with hyperreflexia with brisk reflexes of 3+ at the knees, absent clonus, extensor plantar responses, bilateral pes planus, normal sensory exam, and a markedly spastic gait. No dystonic posture and bradykinesia were noted. Upper limb exam was normal.

##### Twin B clinical presentation

Twin B had normal development until between 5 to 10 years of age when they presented with abnormal gait, which progressed to lower limb spasticity with increasingly frequent falls. Between 10 to 15 years of age, twin B required ankle foot orthosis (AFO) and received occupational therapy and physical therapy on a regular basis. They could carry out most activities of daily living independently, but needed to use a walking aid for ambulation due to frequent falls. They had normal sitting posture, normal bladder function, and was in a regular classroom with grades above average. On physical exam at the age range 10-15, twin B presented with mild scoliosis, mild muscle atrophy spastic tone of lower limbs, hyperreflexia of 3+ at the knees, abnormality in the feet, power was 5/5 proximal, 4-/5 distal with pes cavus of the feet. Plantar responses were equivocal bilaterally. Dystonic posturing and bradykinesia were not detected. Sensory exams including vibration and proprioception was within normal limits. Upper limb exam was normal.

##### Treatment

Both siblings were treated with Sinemet 100/25 (carbidopa/levodopa) at the age range 10-15, and within 6 weeks of starting therapy they showed a remarkable improvement. Formal neurological exam one year after starting the treatment revealed nearly normal muscle tone and strength of the lower extremities, with very mild weakness at the ankles. They were able to ambulate using ankle-foot orthoses and no longer required aid for walking. Due to the initial success of the levodopa trial, we did not perform lumbar puncture to measure catecholamines. Currently aged 15-20, their neurological examination, including deep tendon reflexes and plantar responses, is completely normal, and the patients are able to run and jump.

Both patients had normal creatine kinase (CK) levels, and magnetic resonance imaging (MRI) of the brain and spine revealed no specific abnormalities. Electromyogram (EMG) and nerve conduction velocity (NCV), microarray and metabolic testing were normal.

#### Family 2

Patient C from family 2 is of French-Canadian ancestry, born after a normal pregnancy and delivery without any complications, with normal language and motor skill development until between 5-10 years of age, when they started to have difficulty walking. Two years after the start of symptoms, a possible diagnosis of cerebral palsy was made and 3 and half years after the symptoms started, a diffuse hyperreflexia was noted in lower limbs which was more significant on the right side. In addition, there was a retraction of the right Achilles tendon. When examined between the ages 10-15, their parents reported that they walk with abnormal posture of the toes. They had to stop frequently while walking due to muscle fatigue, and were unable to run. School performance was normal. Symptoms were reported to worsen toward the end of the day or with exercise. Family history was unremarkable. In their physical exam, muscle strength was 5/5 in the upper limbs, 5-/5 in the right leg (more noticeable in proximal muscles). Reflexes were 2+ in upper limbs as well as in the left patella, and 3+ in the right patella. Ankle jerks were brisk bilaterally. They had up-going toes in their right foot associated with 3 beats of clonus. When walking, they tended to drag their right leg slightly. Neither dystonia nor bradykinesia were noted. Over the years, the patient was lost to follow-up, and treatment was not initiated. Following our findings, we were able to locate the patient and suggest that they see a neurologist to consider a trial of levodopa.

### Comparison to previously reported HSP patients with *GCH1* mutations

Table 1 details the main characteristics of the current and previous cases of HSP patients with *GCH1* mutations. Dystonia, the main manifestation of DRD, was not seen in our patients. Also, daily fluctuation, a prominent feature in DRD, which is also one of its diagnostic criteria^37^ was absent in two of our patients. On the other hand, spasticity, which is not usually seen in the autosomal dominant form of DRD (as opposed to autosomal recessive form),^38^ was seen in all three of our patients. Three of the five previously reported HSP patients with *GCH1* mutations also had parkinsonism, whereas our patients did not show signs of bradykinesia, rigidity or tremor.

**Table 1.** Comparison of characteristics of patients in the current study with previously reported HSP patients with *GCH1* mutations.

| Patient | Age<br>range<br>at onset<br>(y) | Age range<br>at<br>diagnosis<br>(y) | First<br>clinical<br>diagnosis | Spastic<br>gait | Spasticity<br>Legs<br>(L/R) | Hyperreflexia<br>(L/R) | Plantar<br>responses<br>(L/R) | Bladder<br>symptoms | Ankle<br>clonus<br>(L/R) | Motor<br>delay | Speech<br>delay/abnormality | Dystonia | Diurnal<br>fluctuations | Parkinsonism | MRI |
| --- | --- | --- | --- | --- | --- | --- | --- | --- | --- | --- | --- | --- | --- | --- | --- |
| 0 | 5-10 | 35-40 | CP | + | +/+ | +/+ | NA | - | - | - | + (dysarthria) | - | + | - | Normal |
| 1.1 | 0-5 | 55-60 | CP | + | +/+ | +/+ | ↑ / ↑ | + | -/- | - | - | + | + | +/- | NA |
| 1.2 | 0-5 | 25-30 | HSP | + | +/+ | +/+ | ↑ / ↓ | — | -/+ | NA | NA | — | ++ | — | Normal |
| 2 | 10-15 | 45-50 | CS<br>HSP | + | +/+ | +/+ | ↓ / ↑ | + | - | NA | NA | ++ | + | + | Normal |
| 3 | 5-10 | 50-55 | HSP | + | +/+ | +/+ | ↑ / = | — | NA | NA | NA | — | + | ++ | NA |
| A* | 0-5 | 10-15 | HSP | + | +/+ | +/+ | ↑ / ↑ | - | - | - | - | — | — | — | Normal |
| B* | 5-10 | 10-15 | HSP | + | +/+ | +/+ | =/= | - | - | - | - | — | — | — | Normal |
| C* | 5-10 | 10-15 | CP | + | +/+ | -/+ | ↑ / ↑ | + | + | + | - | — | + | — | Normal |

Abbreviations: y, year; L, left; R, right; MRI, Magnetic Resonance Imaging; CP, cerebral palsy; NA, not available; HSP, hereditary spastic paraplegia; CS, cervical stenosis; +, present; -, absent; Plantar responses: ↑, extensor; ↓, flexor; =, equivocal.
\* Patients in the current study.

When examining the location of *GCH1* mutations in the current and previously reported cases of HSP, DRD and PD (Supplementary Table 5), there was no significant difference in the location of *GCH1* mutations comparing DRD, HSP and PD. (P=0.740, Pearson Chi-Square).

## Discussion

We describe three HSP patients with likely pathogenic and pathogenic *GCH1* mutations, responsible for ∼0.4% of all HSP patients and families in our cohort with 696 patients from 431 families. Patients A and B are monozygotic twins, who had bilateral spasticity of the lower limbs, spastic gait and hyperreflexia at the knees. In patient C, the main findings were spasticity of the lower limbs, spastic gait, hyperreflexia in the right lower limb, and diurnal fluctuation. Our patients had neither dystonia nor parkinsonism. These findings expand the genotype and phenotype knowledge about *GCH1* mutations in HSP.

The mutation in family 1, p.(Ser77_Leu82del), has not been reported before. However, the mutation in family 2, p.(Val205Glu), has been reported in three studies, two reporting families with typical DRD ^29,30^ and one reporting a patient with atypical DRD with isolated resting leg tremor. ^39^ The two former studies also reported family members who were asymptomatic carriers of the mutation, suggesting an incomplete penetrance, which is also supported by the reported lack of symptoms in the father of the proband who also carried the p.(Val205Glu) mutation. ^29,30^

To date, *GCH1* mutations in HSP have been described in two reports,^16,17^ with the age of onset between 2 and 14 years of age. Our patients had an age range of 0-10 at diagnosis. Two of the patients in the previous report had dystonia, three had parkinsonism, and all five had diurnal fluctuations. These symptoms were absent in our patients, except for diurnal fluctuation, which was reported in one patient. On the other hand, anatomical features (e.g. pes cavus, pes planus, scoliosis) as well as lower extremity weakness were seen in our patients, but have not been previously reported. The differences in phenotypes of the monozygotic twins in the current study (e.g. age at onset, presence of weakness, scoliosis, plantar response, muscle atrophy) may suggest that environment, epigenetics, and stochasticity ^40-42^ could play a role in the presentation of patients with *GCH1* mutations.

Interestingly, *GCH1* mutations which cause DRD, PD and HSP are found in overlapping regions of the protein (figure 2), which decreases the possibility that the location of the mutations alone affects the phenotype. Therefore, other genetic or environmental modifying factors may play a role in disease presentation and the mutation. Other genes have been reported with similarly overlapping phenotypes. For instance, mutations in *ATP13A2* cause Kufor-Rakeb syndrome,^43^ which is an atypical parkinsonism-dystonia disorder. More recently, *ATP13A2* has been reported to also be associated with HSP.^44,45^ *SPG7* is another example of a gene associated with HSP as well as parkinsonism.^46,47^ In addition, *SPG11* mutations have been reported in patients with parkinsonism^48^ and DRD^49^ in addition to HSP.^50^ These reports suggest a clinical-genetic overlap between these disorders, and that screening for mutations in these genes is indicated when the most common genetic causes have been ruled out.

**Figure 2.**
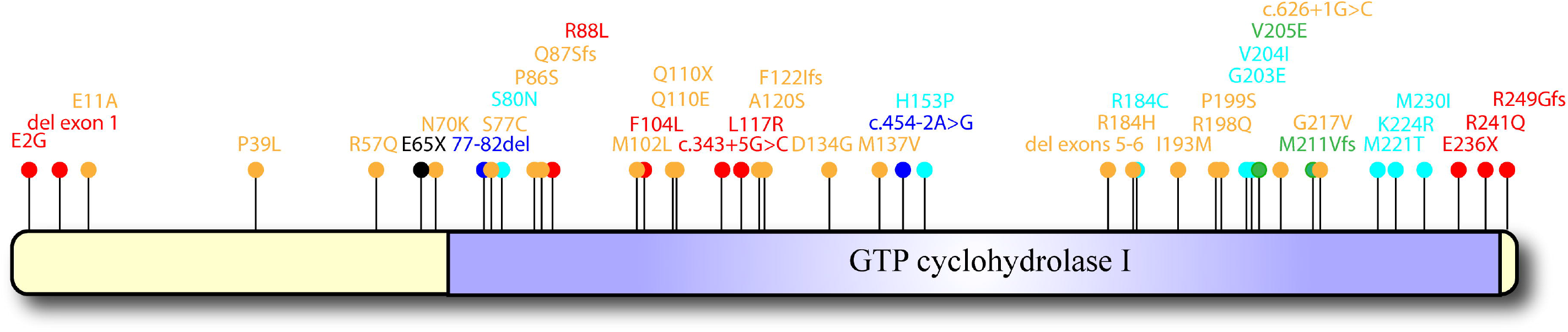
Schematic representation of the location of *GCH1* mutations in hereditary spastic paraplegia (HSP), dopa-responsive dystonia (DRD) and Parkinson’s disease (PD) patients reported so far. The rectangle represents GCH1 protein, and the functional domain, GTP_CycloHydrolase_I, is indicated with vertical lines. Mutations associated with PD, DRD, and HSP are indicated in yellow, red, and dark blue respectively. Circles in green indicate mutations associated with both HSP and DRD, those in light blue with both PD and DRD, and those in black with PD, DRD and HSP.

To conclude, our results provide additional support for the involvement of *GCH1* in HSP and expand the clinical phenotype of *GCH1*-related disorders. We recommend trial of levodopa in HSP patients as well as adding *GCH1* to the sequencing panels of HSP genes to increase their genetic diagnostic yield.

## Data Availability

Data available on request from the authors.

## Acknowledgements

We thank the patients and their families for participating in this study. This study was funded by CIHR Emerging Team Grant, in collaboration with the Canadian Organization for Rare Disorders (CORD), grant number RN127580 – 260005, and by a CIHR Foundation grant granted to GAR. MAE is funded by the Fonds de Recherche du Québec–Santé (FRQS). SV holds a Basic Research Fellowship from Parkinson Canada. JFT holds a Canada Research Chair (Tier 2) in Structural Pharmacology. GAR holds a Canada Research Chair (Tier 1) in Genetics of the Nervous System and the Wilder Pen field Chair in Neurosciences. ZGO is supported by the Fonds de recherche du Québec–Santé Chercheur-Boursier award and is a Parkinson Canada New Investigator awardee. We thank Patrick Dion, Daniel Rochefort, Helene Catoire, and Vessela Zaharieva for their assistance.

## Data availability statement

Data available on request from the authors.

