## Supplementary Table 3 for "*GCH1* mutations in hereditary spastic paraplegia"

**Supplementary table 3** Detailed ACMG criteria. ^1^

| **Mutation** | **ACMG criteria** | | | | | | |
| --- | --- | --- | --- | --- | --- | --- | --- |
|  | **PS3** ^†^ | **PM1** ^‡^ | **PM2**^§^ | **PM4** ^¶^ | **PP2** ^*^ | **PP3** ^**^ | **PP5** ^***^ |
| **p.(Val205Glu)** | Strong | Moderate | Moderate | NA | Supporting | Supporting | Moderate |
| **p.(Ser77_Leu82del)** | NA | Moderate | Moderate | Moderate | NA | Supporting | NA |

Abbreviations: ACMG, American College of Medical Genetics; NA, not applicable.

† Well-established in vitro or in vivo functional studies supportive of a damaging effect on the gene or gene product.

‡ Located in a mutational hot spot and/or critical and well-established functional domain (e.g., active site of an enzyme) without benign variation.

§ Absent from controls (or at extremely low frequency if recessive) in Exome Sequencing Project, 1000 Genomes Project, or Exome Aggregation Consortium.

¶ Protein length changes as a result of in-frame deletions/insertions in a non-repeat region or stop-loss variants.

* Missense variant in a gene that has a low rate of benign missense variation and in which missense variants are a common mechanism of disease.

** Multiple lines of computational evidence support a deleterious effect on the gene or gene product (conservation, evolutionary, splicing impact, etc.)

*** Reputable source recently reports variant as pathogenic, but the evidence is not available to the laboratory to perform an independent evaluation.

1.	Kopanos C, Tsiolkas V, Kouris A, et al. VarSome: the human genomic variant search engine. *Bioinformatics.* 2019;35(11):1978.
