## Supplementary Table 5 for "*GCH1* mutations in hereditary spastic paraplegia"

**Supplementary Table 5** Reported mutations in *GCH1.*

| Reference | Phenotype | | Variant | Location (amino acid 73-248)^†^ |
| --- | --- | --- | --- | --- |
| ^1^ | DRD | | Arg249Gly fs | No |
| ^2^ | DRD | | Arg241Gln | Yes |
| ^3^ | DRD | | Glu236X | Yes |
| ^2^ | PD, DRD | | Met230Ile | Yes |
| ^4^ | PD,DRD | | Lys224Arg | Yes |
| ^4^ | PD, DRD | | Met221Thr | Yes |
| ^2^ | PD | | Gly217Val | Yes |
| ^5^ | DRD,HSP | | Met211Val fsX38 | Yes |
| ^6-8^ | DRD, HSP | | Val205Glu | Yes |
| ^2^ | PD, DRD | | Val204Ile | Yes |
| ^1^ | PD,DRD | | Gly203Glu | Yes |
| ^9^ | PD | | Pro199Ser | Yes |
| ^1^ | PD | | Arg198Gln | Yes |
| ^1^ | PD | | Ile193Met | Yes |
| ^4^ | PD, DRD | | Arg184Cys | Yes |
| ^10^ | PD | | Arg184His | Yes |
| ^1^ | PD,DRD | | His153Pro | Yes |
| ^1^ | PD | | Met137Val | Yes |
| ^2^ | PD | | Asp134Gly | Yes |
| ^1^ | PD | | Phe122fs | Yes |
| ^2^ | PD | | Ala120Ser | Yes |
| ^11^ | DRD | | Leu117Arg | Yes |
| ^2^ | PD, DRD | | Gln110X | Yes |
| ^2^ | PD | | Gln110Glu | Yes |
| ^2^ | DRD | | Phe104Leu | Yes |
| ^1^ | PD | | Met102Leu | Yes |
| ^12^ | DRD | | Arg88Leu | Yes |
| ^1^ | PD | | Gln87Serfs | Yes |
| ^1^ | PD | | Pro86Ser | Yes |
| ^1^ | PD,DRD | | Ser80Asn | Yes |
| ^1^ | PD | | Ser77Cys | Yes |
| ^1^ | PD | | Asn70Lys | No |
| ^1^ | PD,DRD,HSP | | Glu65X | No |
| ^1^ | PD | | Arg57Gln | No |
| ^1^ | PD | | Pro39Leu | No |
| ^1^ | PD | | Glu11Ala | No |
| ^13^ | DRD | | Glu2Gly | No |
| ^2^ | PD | | c.626 + 1G>C | Yes |
| ^14^ | HSP | | c.454-2A>G | Yes |
| ^2^ | DRD | | c.343 + 5G>C | Yes |
| Current paper | HSP | | c.229_246del:p.77_82del | Yes |
| ^15^ | PD | | Deletion of exon 5 and 6 | Yes |
| ^16^ | DRD | | Deletion of exon 1 | Yes |
| ^17^ | PD | Complete deletion of GCH1 | | Yes |

Abbreviations: HSP, Hereditary spastic paraplegia; DRD, Dopa-responsive dystonia; PD, Parkinson’s disease.

† Loss of function mutations were considered to be affecting this domain.

(NM_000161.3)
